# Posture analysis in predicting fall-related injuries during French Navy Special Forces selection course using machine learning: A proof of concept study

**DOI:** 10.1101/2023.07.26.23293231

**Authors:** Charles Verdonk, Anaïs M. Duffaud, Aurélie Longin, Matthieu Bertrand, Fabien Zagnoli, Marion Trousselard, Frédéric Canini

**Affiliations:** French Armed Forces Biomedical Research Institute, Brétigny-sur-Orge, France; Laureate Institute for Brain Research, Tulsa, Oklahoma, United States; VIFASOM (EA 7330 Vigilance Fatigue, Sommeil et Santé Publique), Université de Paris, Paris, France; 125th Medical Unit of Lann Bihoué, Lorient, France; 6th Special Medical Unit of Orléans-Bricy, Bricy, France; Clermont Tonnerre Military Hospital, Department of Neurology, Brest, France; French Military Health Academy, Paris, France

## Abstract

**Introduction:** Injuries induced by falls represent the main cause of failure in the French Navy Special Forces selection course. In the present study we made the assumption that probing the posture might contribute to predicting the risk of fall-related injury at the individual level.

**Methods:** Before the start of the selection course, the postural signals of 99 male soldiers were recorded using static posturography while they were instructed to maintain balance with their eyes closed. The event to be predicted was a fall-related injury during the selection course that resulted in the definitive termination of participation. Following a machine learning methodology, we designed an artificial neural network model to predict the risk of fall-related injury from the descriptors of postural signal.

**Results:** The neural network model successfully predicted with 69.9% accuracy (95% CI=69.3-70.5) the occurrence of a fall-related injury event during the selection course from the selected descriptors of the posture. The area under the curve (AUC) value was 0.731 (95% CI=0.725-0.738), the sensitivity was 56.8% (95% CI=55.2-58.4), and the specificity was 77.7% (95% CI=76.8-0.78.6).

**Conclusion:** If confirmed with a larger sample, these findings suggest that probing the posture using static posturography and machine learning-based analysis might contribute to inform risk assessment of fall-related injury during military training, and could ultimately lead to the development of novel programs for personalized injury prevention in military population.

**KEY MESSAGES:**

- Fall-related injuries are a major concern that leads to failure in the French Navy Special Forces selection course.
- This proof of concept study shows that analyzing the posture with machine learning can predict the risk of fall-related injury at the individual level.
- The findings may prompt the development of novel programs for personalized injury prevention in military settings.

## INTRODUCTION

In the military environment, physical activity and sport play a fundamental role in the development and optimization of combatants’ operational capacity. The benefits of sporting activities last throughout a soldier’s career, as they contribute to the individual’s ability to adapt to operational constraints, and the maintenance of good health, notably with respect to the prevention of chronic, stress-related pathologies (e.g., cardiovascular disease, chronic pain, depression).

However, physical activity and sport are also the cause of acute (e.g., sprains, fractures) and chronic (e.g., tendinopathies) musculoskeletal pathologies. In particular, lower limb trauma is prevalent (15–20%) among young recruits of the French Army, both in the context of operational units,^1,2^ and during selection tests for specialized units (30–45%).^3–5^ In addition to their direct impact on the health of personnel, these injuries also have significant economic and operational costs, due to incapacity.^6^

### Fall injuries during the French Navy Special Forces selection course

In the French Army, a dedicated selection course (*Stage Commando Marine* in French, STAC) aims to select candidates for the French Navy Special Forces (*Commandos Marine* in French). It comprises an initial three-week assessment phase, which is extremely demanding both physically and mentally. Epidemiological data from the assessment phase have shown that medical discharge is the leading cause of failure (70%), and that this is most often the result of acute musculoskeletal trauma (60%), mainly to the lower limbs.^3–5^ In 90% of cases, these injuries are the result of a fall due to a loss of balance.^3^ The physical activities that must be completed during the STAC severely test participants’ balance: candidates must carry heavy loads (a 15 kg combat bag and weapon) for prolonged periods, and complete demanding challenges, with little sleep and regardless of weather conditions. The ‘jungle’ obstacle test is a particularly demanding test; participants must negotiate obstacles up to 5 meters high, with no safety nets or lines, and the course has been reported to be the most frequent source of balance-related injuries.^5^ Overall, the epidemiological data suggest that the candidate’s postural control may play a role in the occurrence of fall-related injuries during the STAC.

### Posturography

Posturography is a tool that is used to characterize postural control.^7,8^ During posturography, the subject stands on a fixed surface, called a stabilometric platform (see Figure S1), and is instructed to maintain balance. From a biomechanical point of view, postural balance in the static standing position is described using the inverted pendulum model: the body moves continuously along a medio-lateral and an antero-posterior axis, both of which originate in the medial malleoli of the ankles.^9^ Both endogenous physiological disturbances (e.g., breathing, the heartbeat) and the physiognomy of the human body (⅔ of the body’s mass is located in the upper ⅔ of the body) mean that the standing position is inherently unstable.^10,11^ Consequently, even when in a static, standing position, postural control must be exercised to maintain balance and avoid falling.

Posturography records a signal that corresponds to the trajectory of the body’s center of pressure (CoP),^7,8^ the latter being considered as the projection on the ground of the person’s center of mass.^11^ Sixteen variables can be extracted from the postural signal that identify differences in postural control between individuals. As these variables combine high *inter-* individual variance with low *intra*-individual variance, linear discriminant analysis can be used to distinguish between individuals.^12^ In the present study, these 16 postural variables are considered as relevant for modeling the risk of fall injury at the individual level.

### Using posture analysis to predict risk of fall injury

Modelling the relationship between the posture and the risk of fall-related injury is complex, not least because it is both non-linear and multifactorial.^13^ Factors are both specific to the individual (not only their postural stability, but also their weight, level of training, etc.) and linked to the environment (weather conditions, the physical challenge, etc.), and interact in unpredictable ways. These issues led us to use a non-linear statistical model to predict the risk of fall injury from an analysis of the postural signal. In particular, we adopted a machine learning-based approach that included the design of an artificial neural network (NN) model for predictive analysis. The NN model has the advantage of being parsimonious—in other words, it requires less experimental data than other non-linear polynomial approaches to model a relationship with a given degree of accuracy.^14^

### Hypothesis

Our hypothesis was that the postural signal, collected before the selection course begins in a static, eyes closed, standing position, and analyzed using a machine learning methodology, would provide information to predict the individual risk of fall injury during the selection course.

## MATERIALS AND METHODS

### Population

The population consisted of 113 STAC male candidates: 53 were recruited during the March 2014 STAC, and 60 during the September 2014 STAC. The only inclusion criteria were being declared medically fit to be a member of the French Navy Special Forces^15^ and being aged under 40. All participants provided written informed consent before participation and were not financially compensated for their involvement in the study. At the end of data collection, 14 participants were excluded from the final analyses due to missing data.

### Procedure

Data were collected at the 190^th^ Medical Unit that provides medical support to the training center for French Navy Special Forces (Lorient). The postural signal was collected before the selection course begins, using the FEETEST 6 platform (TECHNO CONCEPT^®^, France), with the simple instruction to maintain balance for 52 seconds while keeping the eyes closed (Supplementary Methods 1). It should be noted that postural signal recorded under eyes closed condition was assumed to be a better proxy of postural control quality than eyes open condition. Indeed, under eyes closed condition, sensitive pathways of postural control loop only involve bodily signals from the vestibular and somatosensory systems, and cannot benefit from visual information.^10^ The event to be predicted from the postural analysis was a fall injury secondary to a loss of balance that resulted in the definitive termination of participation in the STAC for medical reasons. Information about the event was collected by the staff of the 190^th^ Medical Unit throughout all the selection course, and the causal link between the injury and a fall due to a loss of balance was established retrospectively (Supplementary Methods 2).

### Postural signal processing

The postural signal was processed using custom scripts in Matlab 2021a (Mathworks^®^). We first filtered the postural signal with a 10 Hz high-pass (fourth order Butterworth) filter. Then, the values of the 16 individual-specific descriptors were extracted from the postural signal (Supplementary Methods 3). The 16 descriptors were chosen because they have been characterized in the literature as individual-specific, in other words, they can identify inter-individual differences.^12,16^ In addition to the 16 descriptors, their pairwise products (except products of a descriptor by itself) were computed. Indeed, the product of two descriptors may provide more relevant information than the two descriptors separately, particularly when they are interdependent as is the case for postural descriptors.^14,17^ Thus, a total of 136 candidate postural descriptors were generated, including the 16 descriptors and their 120 pairwise products.

### Predictive model: design and performance assessment

#### Selection of postural descriptors

The purpose of predictor selection is to identify irrelevant or redundant predictors that should be removed from the available dataset before designing the predictive model. Indeed, the inclusion of irrelevant predictors is detrimental to the understanding and performance of the predictive model, while rejecting relevant predictors can be just as bad.^17^ In the present work, the appropriate set of postural descriptors for predicting fall-related injury event was selected among the 136 candidate descriptors in using the forward stepwise regression method,^14,18^ which was implemented via Matlab 2021a (The Mathworks^®^), including the Statistics and Machine Learning toolbox, the Deep Learning toolbox, and custom scripts (Supplementary Methods 4).

#### Data over-sampling

Our dataset was characterized by a number of participants identified as ‘Injured due to a fall’ that was lower than the number of participants identified as ‘Not injured due to a fall’ (see Table 1 in Results), which is known as the “class imbalance problem”.^19^ Learning from a dataset with the class imbalance problem may make the learned predictive model unreliable. In the present study, we addressed the class imbalance problem by implementing a data over-sampling approach, which adds examples to the minority class of the training set, using the SMOTE method^20,21^ (Supplementary Methods 5) implemented in R (R version 4.2.1;^22^ “smotefamily” R package).

**Table 1.** Demographic (age) and biometric (weight and height) variables for the 99 male participants included in the final analyses.

|  | *Variables* |  |  |
| --- | --- | --- | --- |
|  | **Age**<br>**(years)** | **Weight**<br>**(kg)** | **Height**<br>**(cm)** |
| | M ± SD | M ± SD | M ± SD |
| **STAC participants**<br>**(N=99)** | 22 ± 4 | 74 ± 10 | 176 ± 18 |
| **Participants injured due to a fall**<br>**(n=37)** | 22 ± 3 | 73 ± 7 | 177 ± 7 |
| **Participants not injured due to a fall**<br>**(n'=62)** | 22 ± 3 | 75 ± 7 | 179 ± 6 |

N, n and n'=number of participants; kg: kilograms; cm: centimeters; M=mean; SD=standard deviation

#### Data analysis

Our hypothesis was as follows: Is it possible to predict the risk of a participant suffering a fall-related injury from the analysis of his or her posture? The question was addressed using a classification approach where the occurrence of a fall-related injury event during the selection course, as a categorical variable (i.e., presence vs. absence), was predicted from the selected postural descriptors. To this end, we designed a predictive model that was an NN model including one layer with a single hidden neuron (Supplementary Methods 6). NN models are a very popular family of classifiers that are particularly efficient for modeling nonlinear relationships,^14^ such as the relation between physiological signal of posture and fall-related injury.^13^ It should be added that models able to capture linear separability between classes (e.g., Support Vector Machine, Linear Discriminant Analysis) were considered as unsuitable for the purpose of the present classification analysis, because examples from the two classes (i.e., presence vs. absence of fall-related injury) were found as not linearly separable according to the Ho and Kashyap’s method.^14,23^ Demographic and biometric data were analyzed using Mann-Whitney tests and its Bayesian equivalent, the latter facilitating the testing of evidence for the null hypothesis (negative log(BF10) values; Supplementary Methods 7, Table S3), and were performed in JASP version 0.14.1 (https://jasp-stats.org/).

#### Evaluation of prediction performance

The predictive performance of our NN model was assessed by means of 5-fold cross-validation. Fivefold cross-validation entails that the set of available data is split into five disjoint subsets; the model is trained with four subsets (including 80% of available data) and tested in the fifth (the validation set, made of the remaining 20% of available data).^24^ For the performance metrics, including the accuracy, the area under the receiving operating characteristic curve (AUC), the f1-score, the sensitivity (Sens), the specificity (Spec), we estimated 95% confidence intervals based on 100 iterations (Supplementary Methods 8). All steps for data analysis and evaluation of prediction performance were performed using custom scripts in R (tensorflow”, “keras”, “caret”, and “ModelMetrics” packages).

## RESULTS

### Demographic, biometric, and epidemiologic data

Table 1 shows descriptive statistics (age, weight, and height) for the 99 males participants included in the final analyses. Participants injured due to a fall did not differ from uninjured participants in terms of age (*p*=0.92, log(BF10)=−1.46), weight (*p*=0.22, log(BF10) =−0.85) or height (*p*=0.13, log(BF10) =−0.87).

Figure 1 shows the number of participants injured as the result of a fall as a function of time since the start of the STAC.

**Figure 1.**
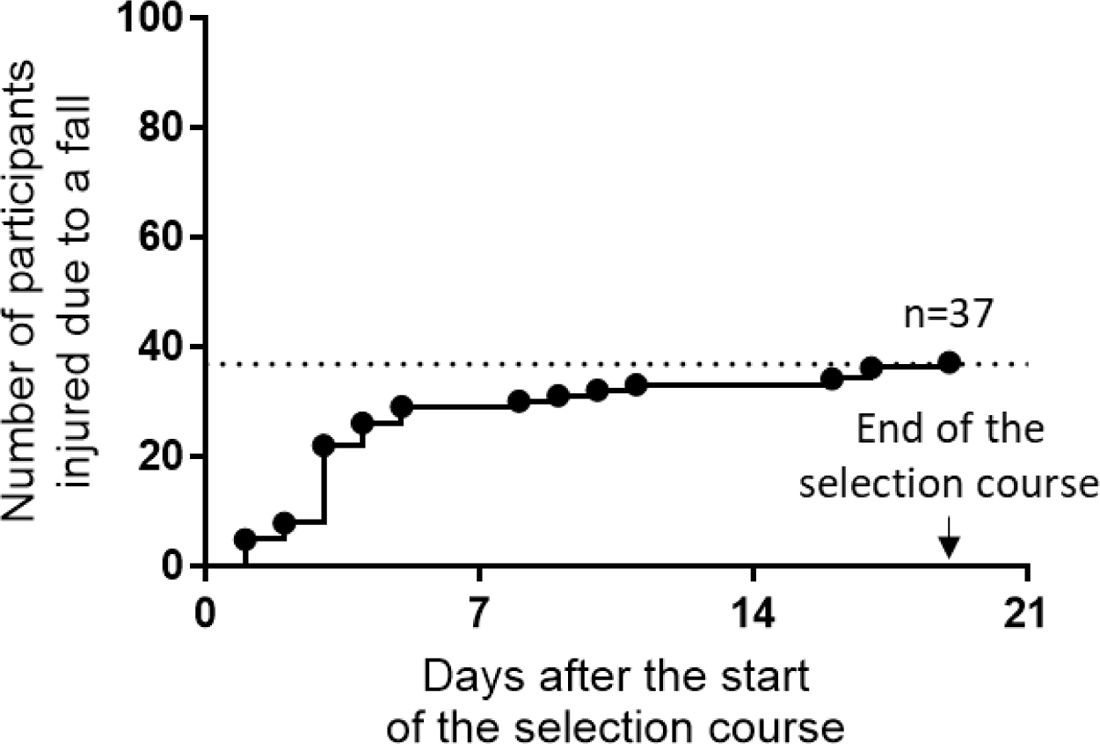
Graphical representation of the number of participants injured as a result of a fall due to loss of balance as a function of the time elapsed since the start of the STAC. Each step corresponds to an event, and the height of the step is proportional to the number of events over the interval.

### Selection of postural descriptors

Table 2 presents the results of the forward stepwise regression method, which identified the postural descriptors that are relevant for predicting fall injury event among the 136 candidate descriptors (including the 16 descriptors and their 120 pairwise products). It should be noted that only the product of two postural descriptors were selected.

**Table 2.** Postural descriptors that were selected for predicting fall injury event among the 136 candidate descriptors (including the 16 descriptors and their 120 pairwise products), based on the forward stepwise regression method. A description including a simplified mathematical definition of each selected descriptor is available in the Supplementary (Supplementary Methods 3).

| Selected postural descriptors |  |
| --- | --- |
| 1. $MP3 \times \log\text{-Alpha-AP}$ | 5. $\log\text{-slope-MP} \times PF95AP$ |
| 2. $Beta\text{-ML} \times \log\text{-Alpha-ML}$ | 6. $\log\text{-MV} \times \log\text{-MV-ML}$ |
| 3. $Beta\text{-AP} \times \log\text{-slope-MP}$ | 7. $\log\text{-MV-AP} \times \log\text{-Power-ML}$ |
| 4. $\log\text{-slope-MP} \times \log\text{-MV-ML}$ | 8. $\log\text{-Power-ML} \times PF95AP$ |

### Prediction performance

Figure 2 and Table 3 show the performance of the NN model for predicting the fall-related injury event during the selection course from the selected postural predictors. Of note, in Table 3, the training set shows the same percentage (50%) of examples identified as ‘Injured due to a fall’ and examples identified as ‘Not injured due to a fall’, as a result of data oversampling step that was implemented to address the class imbalance problem (see Supplementary Methods 5); the validation set includes the same proportion of examples identified as ‘Injured due to a fall’ as the original dataset.

**Figure 2.**
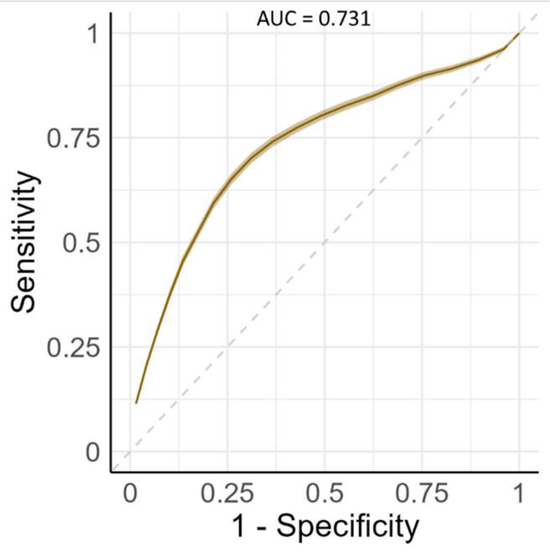
Receiver Operating Characteristic (ROC) curves for the Neural Network model for prediction of the fall-related injury event during the selection course from the selected postural predictors. Error bars of the ROC curves indicate 95% confidence interval for the area under the ROC curve (AUC).

**Table 3.** Performance of the Neural Network model when predicting the fall-related injury event during the selection course from the selected postural predictors (Mean [95% confidence interval]). Results are reported separately for the training set and the validation set.

|  | Performances metrics |  |  |  |  | Percentage of examples belonging to the class 'Injured due to a fall' (%) |
| --- | --- | --- | --- | --- | --- | --- |
|  | AUC | Accuracy | f1-score | Specificity | Sensitivity |  |
| *Training* | 0.805<br>[0.803 - 0.807] | 0.792<br>[0.789 - 0.794] | 0.713<br>[0.710 - 0.716] | 0.856<br>[0.849 - 0.862] | 0.686<br>[0.679 - 0.694] | 50 |
| *Validation* | 0.731<br>[0.725 - 0.738] | 0.699<br>[0.693 - 0.705] | 0.561<br>[0.549 - 0.573] | 0.777<br>[0.768 - 0.786] | 0.568<br>[0.552 - 0.584] | 37 |

AUC: Area under the receiving operating characteristic curve; Injury due to a fall: percentage of participants identified as 'Injured due to a fall'.

## DISCUSSION

Our study analyzed the postural signals of 99 male participants that were recorded before they start the French Navy Special Forces selection course, and we investigated whether postural descriptors can contribute to predict the risk of a fall injury during the selection course. Following a machine learning methodology, a neural network model was designed to predict the risk of a fall injury event from the selected descriptors of postural signal. The main result is that the neural network model was able to predict with 69.9% accuracy the risk of fall-related injury at the individual level. In other words, the analysis of posture allows to correctly predict the risk of fall injury in more than two-thirds of participants of the selection course. This result suggests that it is reasonable to think that analysis of the posture using machine learning-based model is a particularly rich source of information for predicting the risk of fall-related injury. If this result is confirmed with larger sample sizes, and in the context of the other physical activities that armed forces’ personnel undertake, its application would be very interesting for the future of military medicine. Indeed, one of the responsibilities of the military doctor is to preserve the health of the combatant, including the detection of functional weaknesses conducive to injury. A tool that combines postural recording with posturography, and its analysis using a machine learning-based software application, could help the doctor to detect a functional weakness in postural control. Such a device would have the advantage of being quick and easy to use: recording and analysis take just a few minutes. This procedure could supplement other clinical information collected by the doctor. At the present time, the recommendations given to sports medicine practitioners are limited to the use of a clinical test called the Balance Error Scoring System (BESS), which assesses postural control.^25^ In brief, the BESS test assesses three postural control configurations (double leg stance *vs.* single leg stance *vs.* tandem stance) and two types of surfaces (firm *vs.* foam) (see Figure 1 in ^26^). However, this test must be performed by a physician, and the method used to calculate the score has not yet been standardized. Looking ahead, it would be interesting, in the future, to compare results from the BESS test with the analysis of postural signal, both in isolation and in combination, regarding their respective accuracy in predicting the risk of fall-related injury. A medium-term aim is to optimize the clinical-physiological tools that armed services’ doctors can use to detect functional weaknesses in postural control that increase the risk of fall-related injury.

If the functional weakness that contributes to the risk of injury can be identified, appropriate countermeasures and targeted interventions could be implemented to reduce risk.^27^ We hypothesize that improving the individual’s perception of the position of their body in space (i.e., proprioception) could improve postural control, and consequently reduce the risk of fall injury. Interestingly, in the French Army, the Optimization of the Resources of the Armed Forces (ORAF, formerly known as Tactics to Optimized the Potential - TOP) program draws upon cognitive techniques that may be relevant to improving the individual’s perception of their body in space, and includes mental imagery exercises such as a body scan. Regular training and individual practice during physical activities could also help. In this context, the POSITION study (which is currently being conducted in several French Special Forces units) aims to evaluate the effectiveness of the ORAF method in preventing fall-related injuries.^28^ The aim of the POSITION study is to validate or invalidate the usefulness of such a prevention program, based on the ORAF method, in reducing the risk of fall-related injury during the physical activities undertaken by military personnel.

In the present study, the design of our predictive model included the selection of postural descriptors, based on their relevance in predicting fall injury risk. The vast majority of the selected descriptors relate to the CoP velocity profile. It must be acknowledged that the interpretation of these results remains difficult. What is the functional (biomechanical) significance of the selected descriptors in terms of postural control? In other words, how is postural control associated with the risk of falling, characterized from a biomechanical point of view? At the present time, the available data do not offer a simple explanation of what characterizes the postural control of an individual who is at risk of fall injury.

Our study modeled the risk of fall injury based solely on a postural analysis. This reductionist approach, which was intended to simplify the development of our machine learning algorithm, does not take into account the (often multifactorial) nature of the injuries that can occur during sporting activities.^13^ In the specific context of the French Navy Special Forces selection course, other factors, notably fatigue and anxiety, could play an important role. For example, it has been shown that participants with a sleep debt (between three and 20 hours) are five times more likely to suffer a fall-related injury during the selection course.^3^ Nevertheless, the analysis of the timing of the occurrence of injuries suggests that fatigue may not be the predominant risk factor. Indeed, the majority of injuries (75%) occur during the first two weeks of the course, rather than the last week, when fatigue levels are highest.^3–5^ At the same time, anxiety levels are probably very high due to both the context and the psychophysiological demands imposed by the selection course. Earlier work shows that anxiety increases among STAC participants.^4^ The role of anxiety in fall injuries is a particularly important avenue of further study, as the neurophysiological systems involved in postural control are influenced by stress response regulation mechanisms.^29^ For example, the excitability of the postural myotatic reflex, at the level of the spinal cord, is modulated by subcortical structures (notably the periaqueductal gray matter and the amygdala) that are activated during the stress response. It is interesting to note that body awareness plays an important role in regulating the stress response, as it contributes to the identification, evaluation, and modulation of the body’s internal physiological state (also known as interoception), which characterizes the neurobiological response to stress response and the related emotional feeling of anxiety.^30,31^ Thus, body awareness could influence the occurrence of fall injuries either directly, as a central player in the postural control loop, or indirectly, by acting on non-postural pathways that affect posture, such as the stress axis. Future work should focus on integrating these two dimensions (postural and interoceptive) in order to characterize the mechanisms that play a role in the prevention of fall injuries.

## Conclusion

This study demonstrates the potential of posture analysis in predicting the risk of fall-related injuries during the French Navy Special Forces selection course. The findings provide early evidence that static posturography in combination with a machine learning-based predictive model can be a valuable tool for assessing individual risk and potentially preventing injuries in military training. The application of this approach, if confirmed with larger sample sizes and in broader contexts, could significantly impact military medicine by aiding in the detection of functional weaknesses and guiding personalized injury prevention programs.

## Data Availability

We do not have permission to deposit military data in a public repository, but the French Military Health Service at can be approached for any queries related to the data used in this paper.

## Acknowledgments

We would like to thank all the staff of the 190^th^ Medical Unit, which provides medical support to the French Navy Special Forces training center (Lorient), for their help with data collection. We would also like to thank the staff of the Neurophysiology of Stress Unit at the French Armed Forces Biomedical Research Institute for their help with data processing and analysis.

## Contributors

FC, MT, and FZ conceptualized the research question. AL, MB, and AD conducted data collection. CV performed data analysis and drafted the original version of the manuscript. FC, MT, AD, and FZ contributed to edit the original version of the manuscript. All authors approved the final version of the manuscript for submission.

## Ethical considerations

The data presented in this article were collected in the context of a study that was approved by the regional ethics committee (*Comité de Protection des Personnes*, CPP OUEST VI, n° 2014PPRC04), in accordance with the principles of the Declaration of Helsinki (1964) and its subsequent amendments.

## Authors’ notes

This study is part of a project supported by the French Military Health Service. The opinions or assertions expressed herein are the private views of the authors and are not to be considered as official or as reflecting the views of the French Military Health Service and the Laureate Institute for Brain Research.

## Data accessibility

The data and materials are currently private for peer review.

## SUPPLEMENTARY CONTENT

### Supplementary Methods 1. Postural signal measurement

The FEETEST 6 platform comprises four small independent platforms that measure the ground reaction force (linked to the action of the body weight) with respect to the heel and metatarsals, for each foot separately. Mean reaction forces along antero-posterior and medio-lateral axes give the position of the body’s center of pressure (CoP). Hence, the CoP corresponds to the projection, onto the ground, of the sum of the pressures exerted on the various parts of the body that are in contact with the ground. The postural signal was sampled at 40 Hz and transmitted to a software package (PostureWin 4^©^) via a USB connection (Figure S1). Due to technical specifications of the FEETEST 6 platform, the recording of postural signal lasted 52 s. The participants were instructed to stand quietly with eyes closed, and with their arms hanging at their sides and head in a normal forward-facing position.

**Figure S1.**
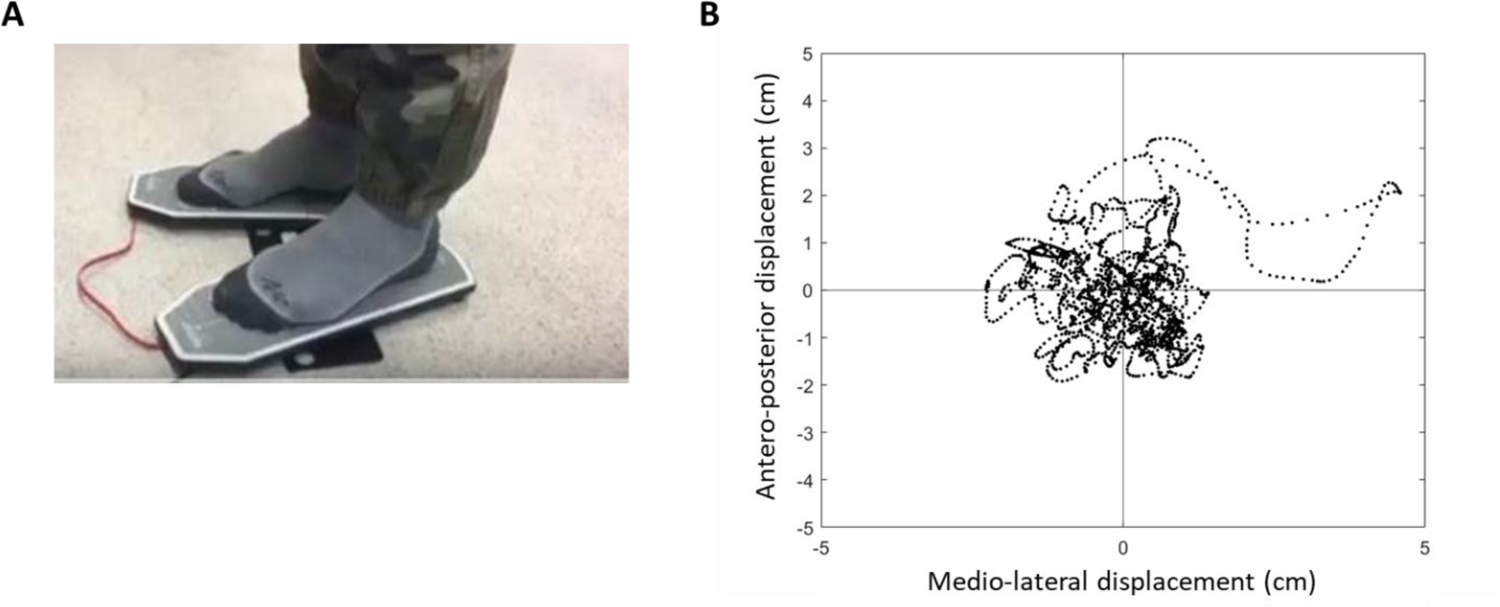
**(A)** The stabilometric platform. **(B)** A sample postural signal collected after filtering and normalization. This signal corresponds to the position of the body’s center of pressure (CoP) along medio-lateral (*x*) and antero-posterior (*y*) axes. The origin of these axes (0, 0) represents the midpoint of the distance between the two medial malleoli. cm: centimeters;

### Supplementary Methods 2. Definition of fall-related injury

The event to be predicted from the postural analysis was a fall injury secondary to a loss of balance that resulted in the definitive termination of participation in the STAC for medical reasons. The causal link between the injury and a fall due to a loss of balance was established retrospectively. Specifically, an initial patient history and diagnosis was carried out by the doctor responsible for providing healthcare support during STAC activities, then six other doctors assessed whether or not the injury was related to a fall due to a loss of balance. Thus, each injury was the subject of a diagnosis, and six assessments of the causal link with a fall due to a loss of balance. A causal link was considered to be valid if at least four doctors assessed the injury as being the consequence of the fall, and the participant was identified as ‘Injured due to a fall’ (membership of the class being equal to 1). In other cases, the injury was not considered as secondary to a fall, and the participant was identified as ‘Not injured due to a fall’, along with participants who did not suffer any injury (membership of the class being equal to 0).

### Supplementary Methods 3. Descriptors of the postural signal

The Table S1 summarizes the 16 individual-specific descriptors that were extracted from the postural signal. The 16 descriptors were chosen because they have been characterized in the literature as individual-specific, in other words, they can identify inter-individual differences.^12,16^ To ensure a similar order of magnitude, they were normalized by subtracting the mean and dividing by the standard deviation obtained at group level (Table S2).

**Table S1.** The list of the 16 descriptors that were extracted from the postural signal to characterize time-series of the center of pressure. A detailed description of the postural features, including their computation is available in the *Supplementary information* of Verdonk et al (2022).^16^.

|  | **Descriptor name** | **Description** |
| --- | --- | --- |
| 1. | MP3 | Mean peak value on sway-density curve at $R = 3$ |
| 2. | Mean-AP | Mean position of sway on AP axis |
| 3. | Mean-ML | Mean position of sway on ML axis |
| 4. | Zero-cross-V-AP | The number of zero crosses of low-pass filtered CoP velocity on AP axis |
| 5. | Beta-ML | Scale parameter of Gamma distribution fitted to the duration of mean CoP velocity crosses on ML axis |
| 6. | log-Alpha-ML | Log of shape parameter of Gamma distribution fitted to the duration of mean CoP velocity crosses on ML axis |
| 7. | log-Alpha-AP | Log of shape parameter of Gamma distribution fitted to the duration of mean CoP velocity crosses on AP axis |
| 8. | Beta-AP | Scale parameter of Gamma distribution fitted to the duration of mean CoP velocity crosses on AP axis |
| 9. | log-slope-MP | Log of slope of the line obtained by linear regression of mean peak values on sway-density curve vs. $R$ from 2 mm to 5 mm |
| 10. | log-LNG | Log of total path length of CoP trajectory on the horizontal plane |
| 11. | log-MV | Log of mean CoP velocity |
| 12. | log-MV-ML | Log of mean CoP velocity on ML axis |
| 13. | log-MV-AP | Log of mean CoP velocity on AP axis |
| 14. | log-Power | Log of total power of CoP |
| 15. | log-Power-ML | Log of total power of CoP on ML axis |
| 16. | PF95AP | 95% power frequency of CoP |

$R$ : radius of circle centered at the current CoP point (in mm); AP: anterior-posterior; ML : medio-lateral; CoP: center of pressure;

**Table S2.** Means and standard deviations prior to standardization of the 16 descriptors extracted from the postural signal.

| Postural features (unit) | M ± SD |
| --- | --- |
| MP3 (sec) | 2.08 ± 0.84 |
| Mean-AP (mm) | 47.78 ± 18.26 |
| Mean-ML (mm) | -1.86 ± 8.80 |
| Zero-cross-V-AP (a.u.) | 113.07 ± 14.92 |
| Beta-ML (a.u.) | 0.13 ± 0.04 |
| log-Alpha-ML (a.u.) | 0.28 ± 0.18 |
| log-Alpha-AP (a.u.) | 0.31 ± 0.16 |
| Beta-AP (a.u.) | 0.16 ± 0.05 |
| log-slope-MP (a.u.) | 0.22 ± 0.60 |
| log-LNG (a.u.) | 6.16 ± 0.30 |
| log-MV (a.u.) | 2.22 ± 0.30 |
| log-MV-ML (a.u.) | 1.34 ± 0.39 |
| log-MV-AP (a.u.) | 2.02 ± 0.29 |
| log-Power (a.u.) | 3.44 ± 1.12 |
| log-Power-ML (a.u.) | 2.82 ± 0.80 |
| PF95AP (a.u.) | 1.02 ± 0.28 |

M: mean; SD: standard deviation; sec: seconds; mm: millimeters; a.u.: arbitrary unit

### Supplementary Methods 4. Selection of postural descriptors

We selected the appropriate set of postural descriptors for predicting fall-related injury event in using the forward stepwise regression method, which has been extensively used in the medical literature and has been shown to achieve the best performance for predictor selection.^18^ Briefly, the method consists in designing different regression models whose independent variables are subsets of the predictors. All the models are compared to a reference model, which specifically includes all of the predictor variables as inputs, and the best model according to a preset criterion is selected.^14^ In the present work, we used the Akaike’s information criterion (AIC) as a decision criterion.

### Supplementary Methods 5. Data over-sampling

Our dataset was characterized by a number of participants identified as ‘Injured due to a fall’ that was lower than the number of participants identified as ‘Not injured due to a fall’ (see Table 1 in the main text). In other words, there was a skew existing between the two different classes of participants regarding the number of examples, which is also known as the “class imbalance problem”.^19^ Learning from a dataset with the class imbalance problem will make the learned predictive model unreliable, because it is more concerned with identifying the majority class (here, the participants identified as ‘Not injured due to a fall’) correctly and ignoring the minority class (here, the participants identified as ‘Injured due to a fall’). We addressed the class imbalance problem by adding examples to the minority class of the training set (data over-sampling) using the SMOTE method (“smotefamily” R package). The main idea behind the method is to create new minority class examples by interpolating several minority class examples that lie together. Specifically, SMOTE created new synthetic examples by randomly selecting a minority class example and generating a new example from random interpolation with one of the k (k = 5) nearest neighbors of the initial example.^20,21^

### Supplementary Methods 6. Classification analysis using a neural network

#### Classification analysis

Our hypothesis was as follows: Is it possible to predict the risk of a participant suffering a fall-related injury as a result of a loss of balance from an analysis of his or her posture? The question was addressed using a classification approach: our predictive model (defined below) assigned each participant to one of two classes, ‘Injured due to a fall’ or ‘Not injured due to a fall’, based on the selected postural descriptors. The classification was divided into two steps: (*i*) an *a posteriori* estimation of the probability of the participant belonging to the ‘Injured due to a fall’ class, based on his or her postural descriptors; and (*ii*) the assignation of each participant to one of the two classes according to the following rule: if the *a posteriori* probability is greater than 50%, the participant is assigned to the ‘Injured due to a fall’ class, otherwise he or she is assigned to the ‘Not injured due to a fall’ class.

**Figure S2.**
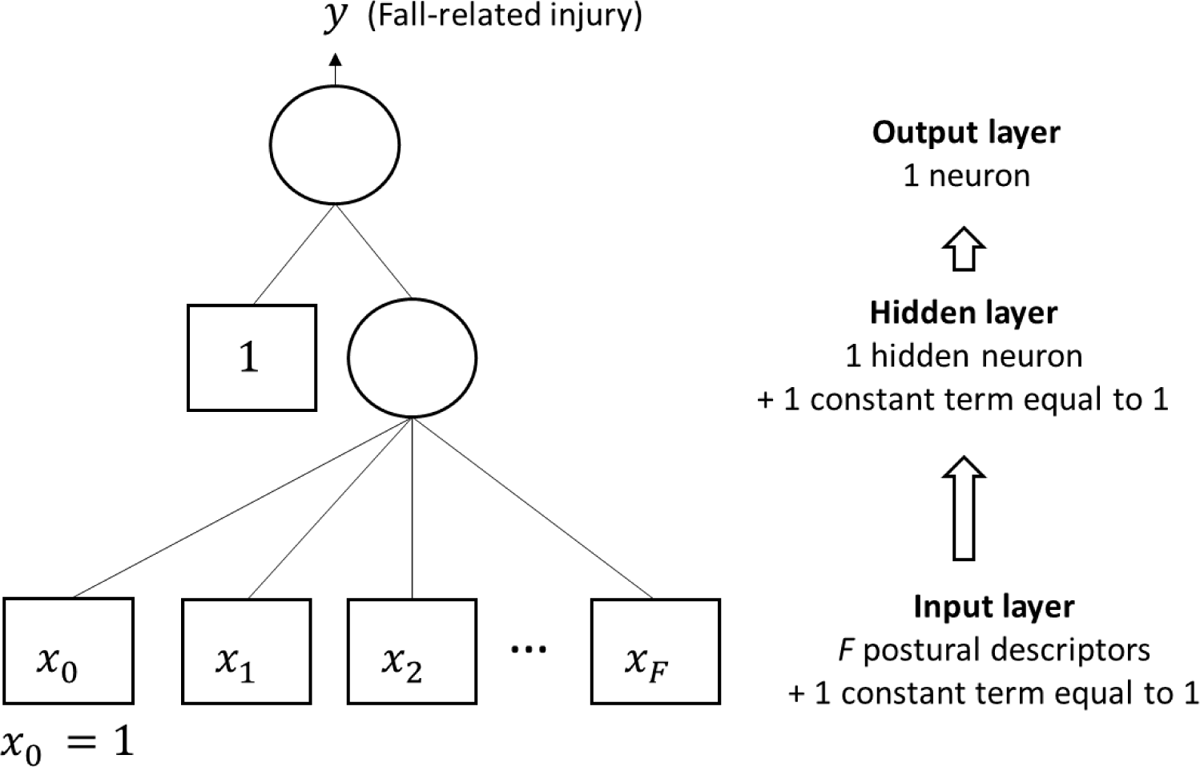
Graphical representation of the neural network model. Information propagates from the bottom (input) to the top (output) layer, via a single intermediate (hidden) layer that includes a single neuron. In our study, input variables were the postural descriptors (*x*_*j*_*j=1*, …, F) and a constant *x*_0_equal to 1, and the output variable *y* was the probability of belonging to the class *Injured due to a fall*.

#### Neural network

The classification analysis was implemented using an artificial neural network. A neural network is a statistical model that combines non-linear functions known as ‘hidden’ neurons. In the present work, we used the simplest form of neural network, called a perceptron, in which one layer includes a single hidden neuron. A graphical representation of our neural network is shown in Figure S2. Input variables were the postural descriptors (*x*_*j*_, *j*=1, …, F) and a constant equal to 1. The single neuron in the hidden layer was a non-linear function (the hyperbolic tangent) of the linear combination of postural descriptors. The output variable, denoted *y*, was the probability of belonging to the ‘Injured due to a fall’ class, calculated as the logistical transformation of the output of the hidden layer neuron. The values for the NN hyperparameters learning rate and number of epochs were 0.14 and 95, respectively.

### Supplementary Methods 7. Bayesian statistical analysis

**Table S3.**
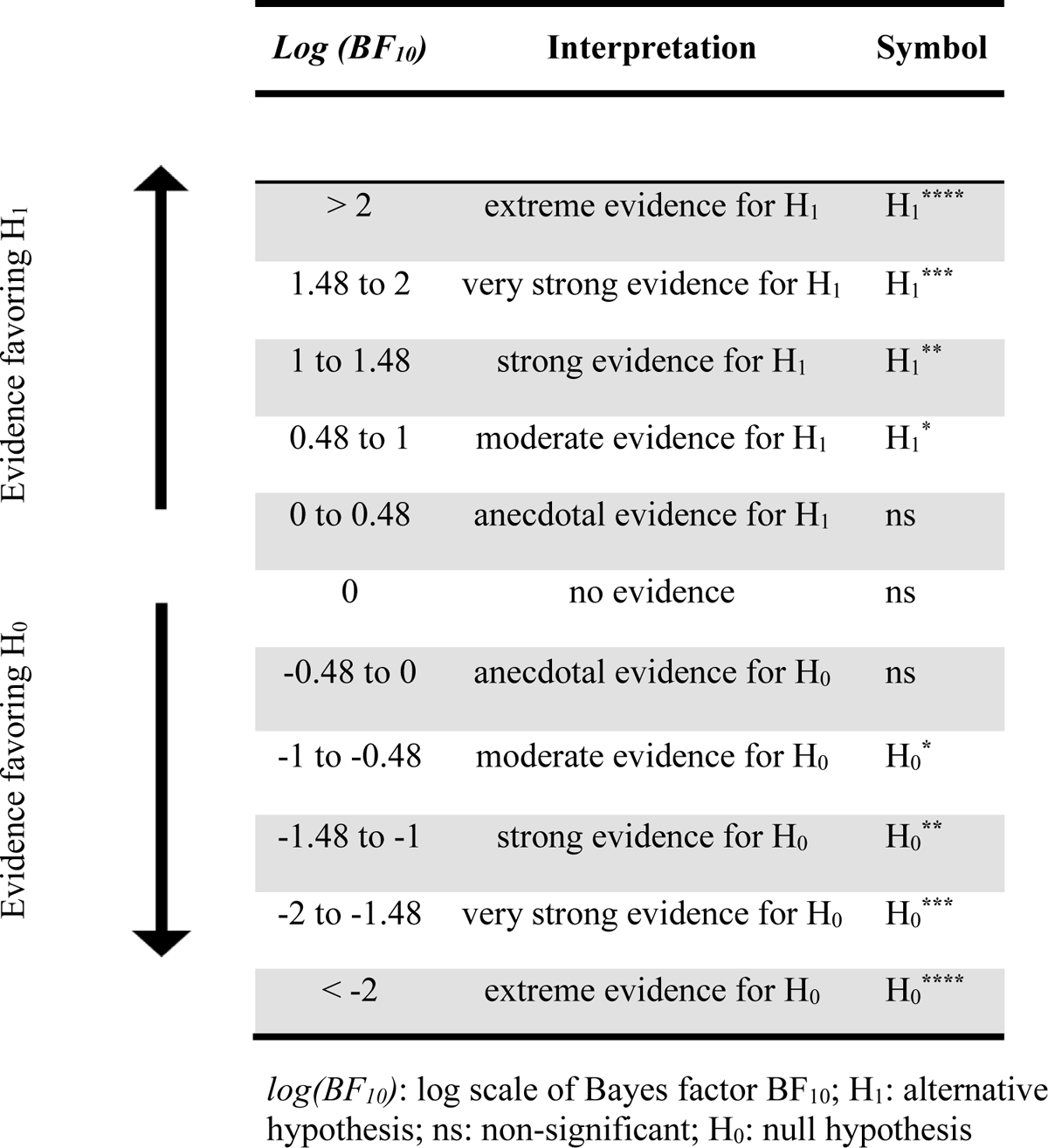
A descriptive and approximate classification scheme for the interpretation of the log scale of Bayes factor (Log (BF10), adapted from Jeffreys, 196.^32^.

### Supplementary Methods 8. Evaluation of prediction performance

To evaluate the performance of our predictive model, the following three indicators were computed:

#### 1 Accuracy

The accuracy represents the percentage of examples that are correctly classified:

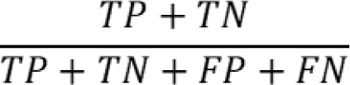

where TP, TN, FP, and FN represent true positive, true negative, false positive, and false negative respectively.

#### 2 f1-score

The f1-score is designed to provide a better metric of model performance for imbalanced datasets, as was the case for the validation sets. The f1-score is based on the harmonic average of precision and recall, which are computed as follows:

- Precision The precision represents the percentage of examples identified as ‘Injured due to a fall’ among the examples classified as ‘Injured due to a fall’ by the predictive model:

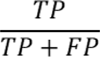
- Recall The recall (also known as sensitivity) represents the percentage of examples correctly classified as ‘Injured due to a fall’ by the predictive model among the examples that are identified as ‘Injured due to a fall’:

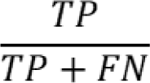

Then, the f1-score is computed as follows:

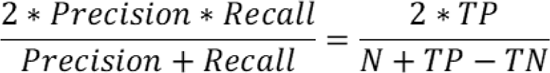

where N is the total number of examples

#### 3 ) Area under the receiving operating characteristic curve (AUC)

The Receiver Operating Characteristic (ROC) curve depicts graphically the relation between sensitivity (or recall) and (1 − specificity) (where specificity is the percentage of examples identified as ‘Not injured due to a fall’ that are (incorrectly) classified as ‘Injured due to a fall’ by the predictive model), at various thresholds. Each threshold value represents decision boundary, within the range [0, 1], for predicting whether an example belongs to the class ‘Injured due to a fall’: for instance, an example is classified as ‘Injured due to a fall’ if the corresponding output of the model is above threshold, otherwise it is considered as ‘Not injured due to a fall’. The area under the ROC curve (AUC) quantifies the ability of the model to correctly assign an example to the class ‘Injured due to a fall’. A value of 1 denotes perfect classification performance, whereas a value below 0.5 means that the model does not perform better than chance level.

